# Covid-19 prevalence estimation by random sampling in the wider population - Optimal sample pooling under varying assumptions about true prevalence

**DOI:** 10.1101/2020.05.05.20075275

**Authors:** Ola Brynildsrud

## Abstract

The number of confirmed Covid-19 cases in a population is used as a coarse measurement for the burden of disease. However, this number depends heavily on the sampling intensity and the various test criteria used in different jurisdictions. A wide range of sources indicate that a large fraction of cases go undetected. Estimates of the true prevalence of Covid-19 can be made by random sampling in the wider population. Here we use simulations to explore confidence intervals of prevalence estimates under different sampling intensities and degrees of sample pooling.

## INTRODUCTION

It is widely accepted that a large fraction of Covid-19 cases go undetected. A crude measure of population prevalence is the fraction of positive tests at any given date. However, this is subject to large ascertainment bias since tests are typically only ordered from symptomatic cases, whereas a large proportion of infected might show little to no symptoms [1,2]. Non-symptomatic infections can still shed the SARS-CoV-2 virus and are therefore detectable by PCR-based tests. It is therefore possible to test randomly selected individuals to estimate the true disease prevalence in a population. However, if the disease prevalence is low, very little information is garnered from each individual test. Under such situations it can be advantageous to pool individual patient specimens into a single sample [3]. Pooling strategies can be efficient to increase the test capacity and are less wasteful with ingredients required for the reverse transcriptase PCR test.

## METHODS

We simulated the effect sample pooling had on prevalence estimates under five different settings for true prevalence *p*. We started by generating a population of 500,000 individuals and then let each individual have *p* probability of being infected at sampling time. The number of patient samples collected from the population is denoted by *n*, and the number of patient samples that are pooled into a single well is denoted by *k*. The total number of pools are thus 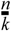, which we call *m*. The number of positive pools in an experiment is termed *x*. We estimate 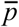 at each parameter combination by replicating the experiment 10,000 times and report here the 2.5% and 97.5% quantiles of the distribution of 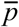.

Explored parameter options:

*p* ∈ {0.001, 0.003, 0.01, 0.03, 1.0}
*n* ∈ {200, 500, 1000, 1500, 2000, 3000, 5000}
*k* ∈ {1, 3, 5, 7, 10, 13, 15, 20, 25, 30, 40, 50, 70, 100, 200}

We considered the specificity (θ) of a PCR-based test to be 1.0, but include simulations with the value set to 0.99. Test sensitivity depends on a range of uncontrollable factors such as virus quantity, sample type, time from sampling, laboratory standard and the skill of personnel [4]. There have also been reports of it varying with pooling level [5]. For the purposes of this study, we fixed the sensitivity (η) at 0.95, irrespective of the level of pooling. This estimate is rather low, which would suggest that we are somewhat overestimating the uncertainty of 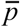. However, since it is possible that tests will be carried out under suboptimal and non-standardized conditions we prefer to err on the side of caution.

The formula of Cowling *et al*., 1999 [6] was used to calculate 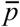 from a single sample:

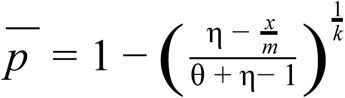

Note that the number of positive pools, *x*, can be approximated in infinite populations as a stochastic variable subject to a binomial distribution with parameters *m* and *P*, where the latter is the probability that a single pool will test positive. A positive pool can arise from two different processes: There can be one or more true positive samples in the pool and they are detected, or there can be no true positive samples in the pool, but the test gives a false positive result:

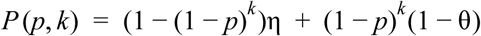

Closer inspection of the above formula reveals something disheartening: In low-prevalence scenarios, and for typical values of test sensitivity and specificity, most positive test results will be false positives.

### Freedom from disease

Pooled sampling can also be used to efficiently assert freedom from disease with a certain probability. If the population is free from the disease, then we find no true positive specimen in our sampling. The question then becomes how many samples we need to take from a population with prevalence *p* to ensure that the probability of sampling at least one single positive patient is *α* or higher. We calculate this number using the formula of Christensen and Gardner, 2000 [7]:

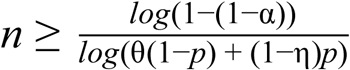

For tests with perfect specificity, we do not have to worry about false positives, and if any pools come out as positive we classify the population as not free from disease. The formula of Christensen and Gardner can be expanded to the case with pooled sampling:

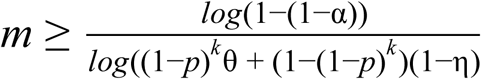

## RESULTS AND DISCUSSION

### Estimates of prevalence

In the following figures, we use simulations to calculate the central 95% estimates of 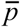. Note that in some parameter combinations, the wavy shape of some curves, particularly for the lower sample sizes 200 and 500, indicates that the precision can actually increase with higher levels of pooling. This is partly due to stochasticity of these results and partly to the discrete nature of each estimate of 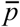. That is, 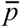 is not continuous and for limited pool sizes miniscule changes in the number of positive pools can affect 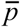 quite a bit.

For example, if you take 200 samples and go with a pool size of 100, there are only three potential outcomes: Both pools are negative, in which case you believe the prevalence is 0; One pool is positive and the other negative, in which case you estimate 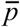 as approximately 0.007 if the test sensitivity is 0.95; Or both pools are positive, in which case the formula of Cowling *et al*. does not provide an answer because the fraction of positive pools is higher than the test sensitivity. This formula is only intended to be used when the fraction of positive pools is much lower than the test sensitivity.

In general, very high levels of pooling are not appropriate since, depending on the true prevalence, the probability that every single pool has at least one positive sample approaches 1. On the other hand, in low prevalence settings, it can be appropriate to pool hundreds of samples, but the total number of samples required to get a precise estimate of the prevalence is much higher. Thus, decisions about the level of pooling need to be informed by the prior assumptions about prevalence in the population, and there is a prevalence-dependent sweet spot to be found in the tradeoff between precision and workload.

**Fig. 1.**
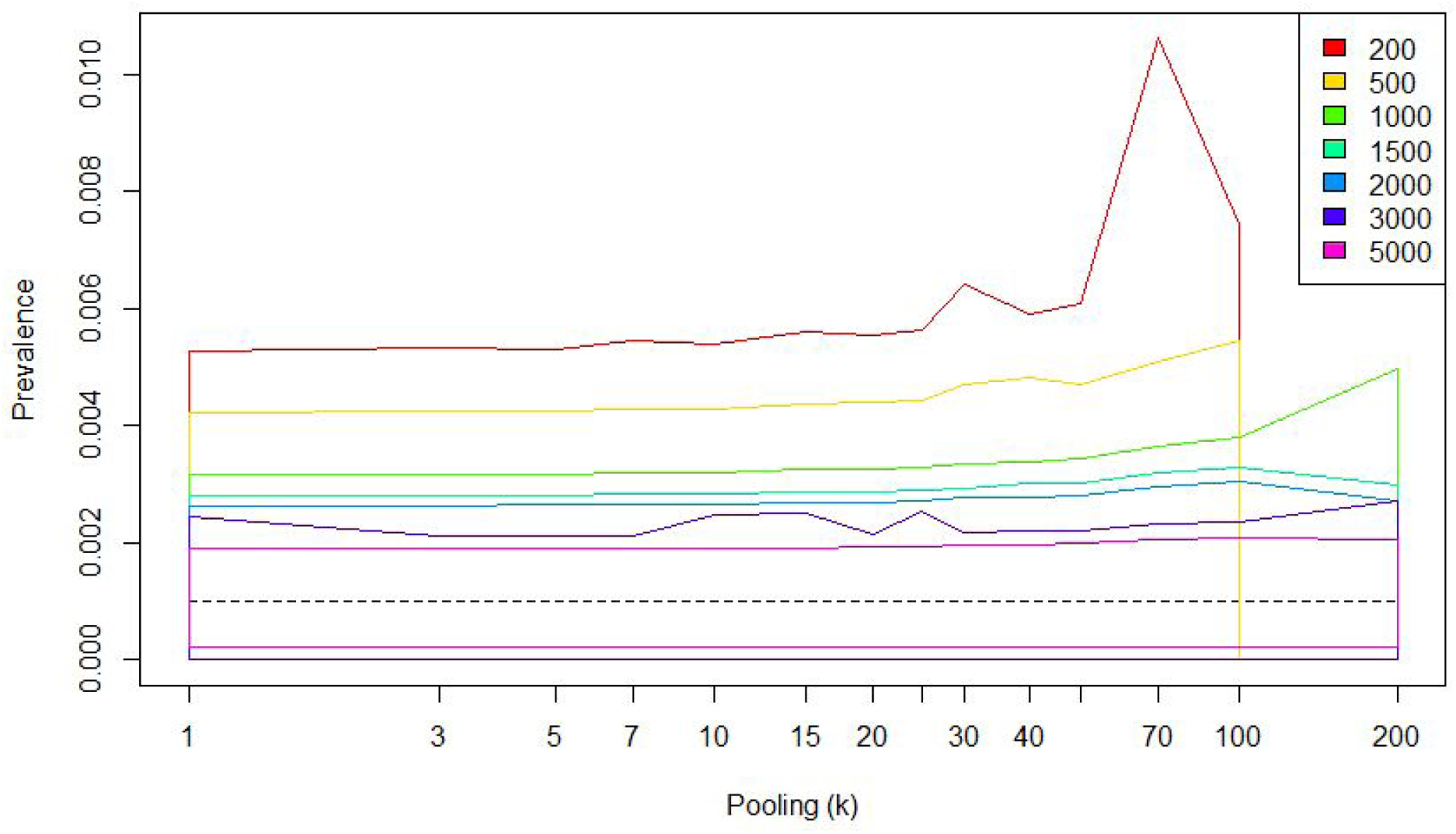
True prevalence = 0.1%:

**Fig. 2.**
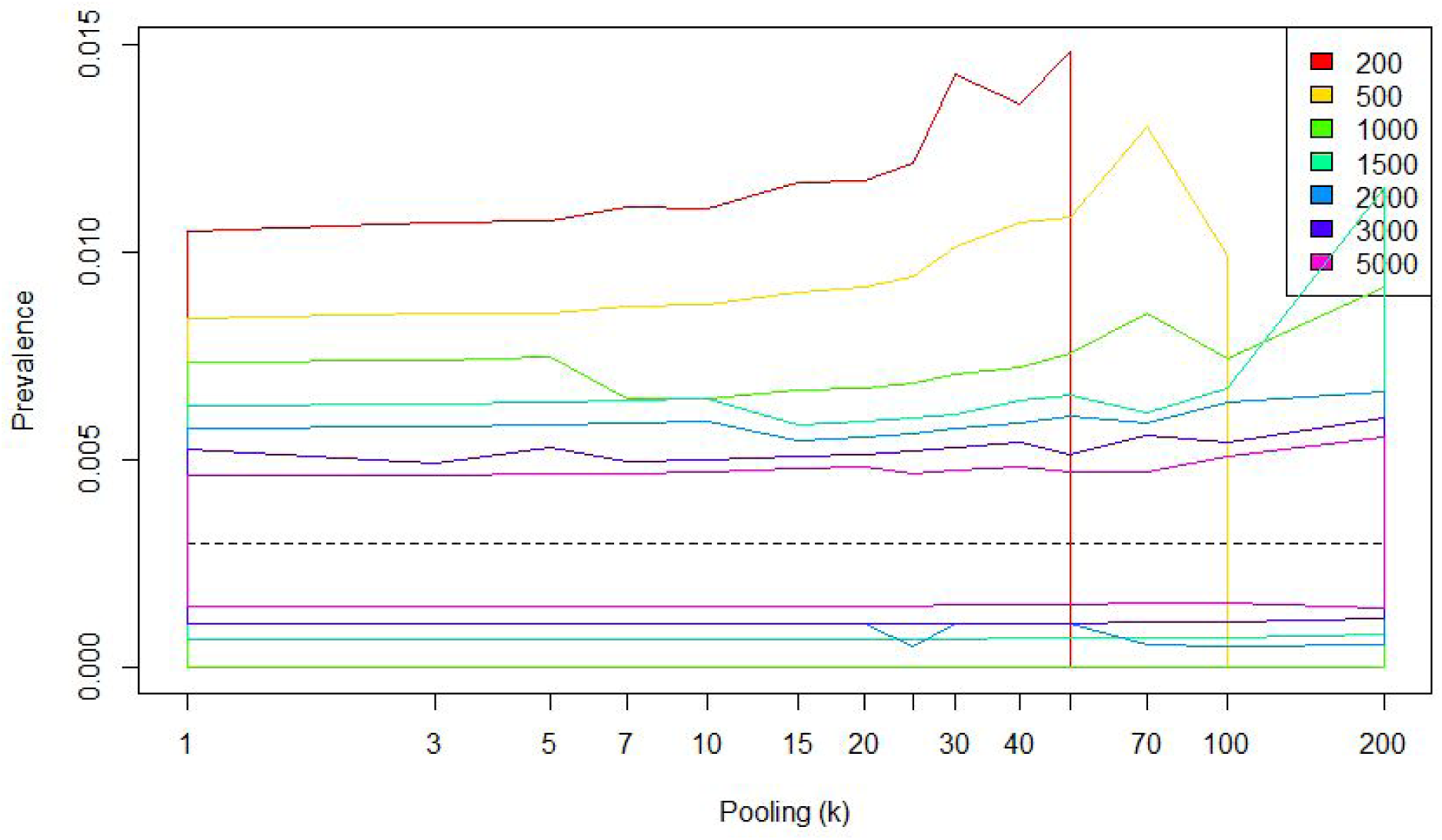
True prevalence = 0.3%:

**Fig. 3.**
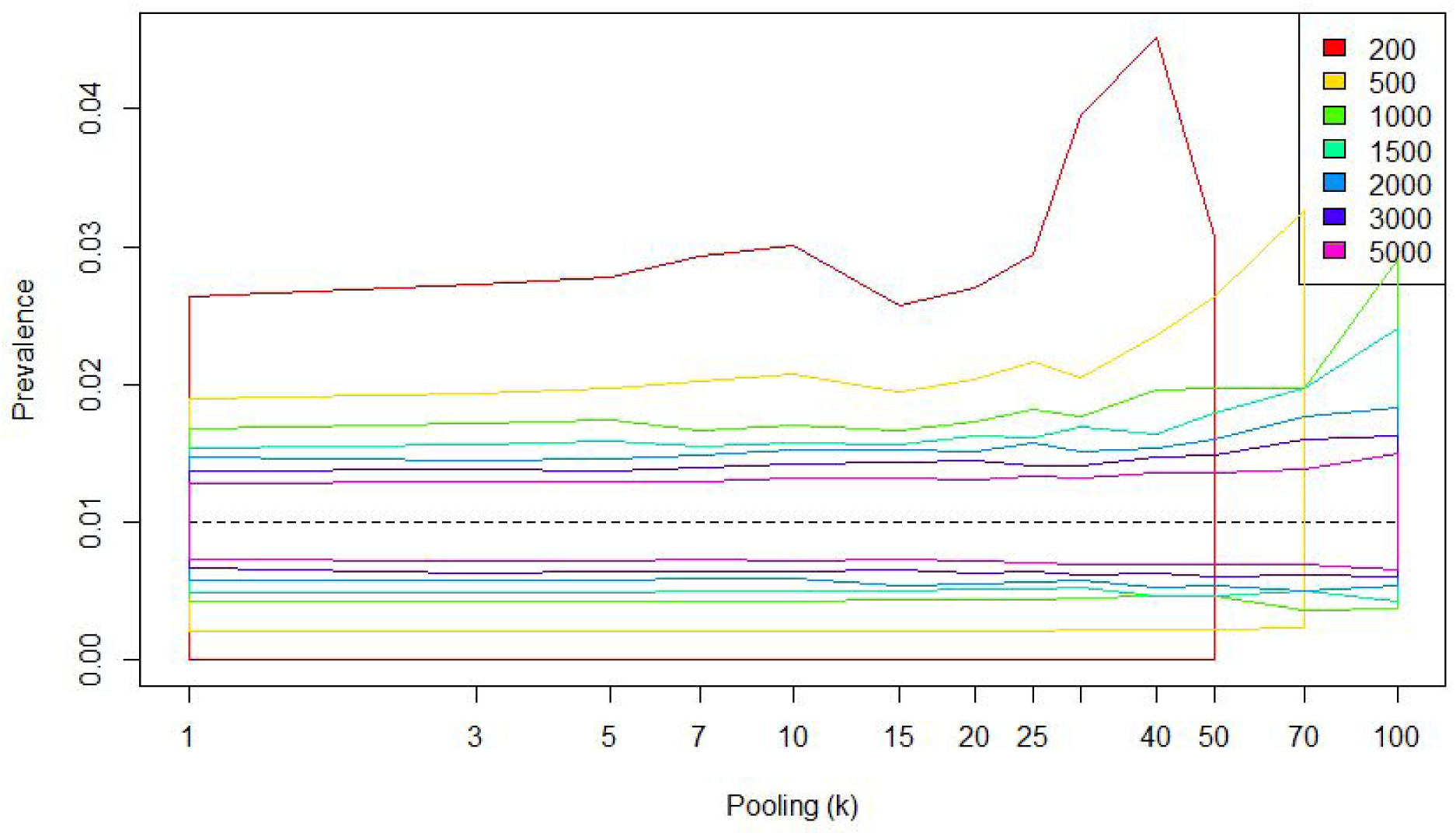
True prevalence = 1%.

**Fig. 4.**
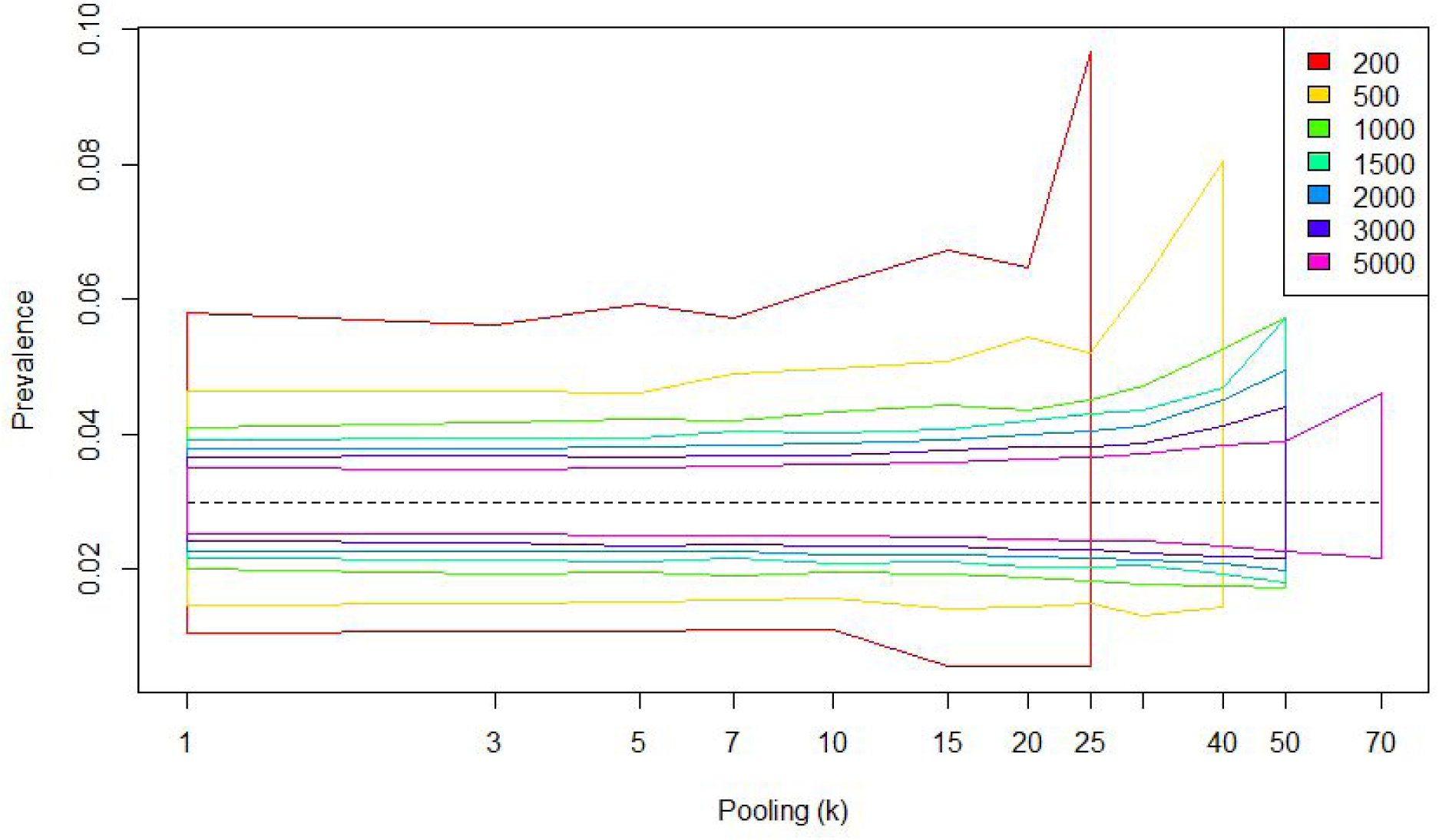
True prevalence = 3%:

**Fig. 5.**
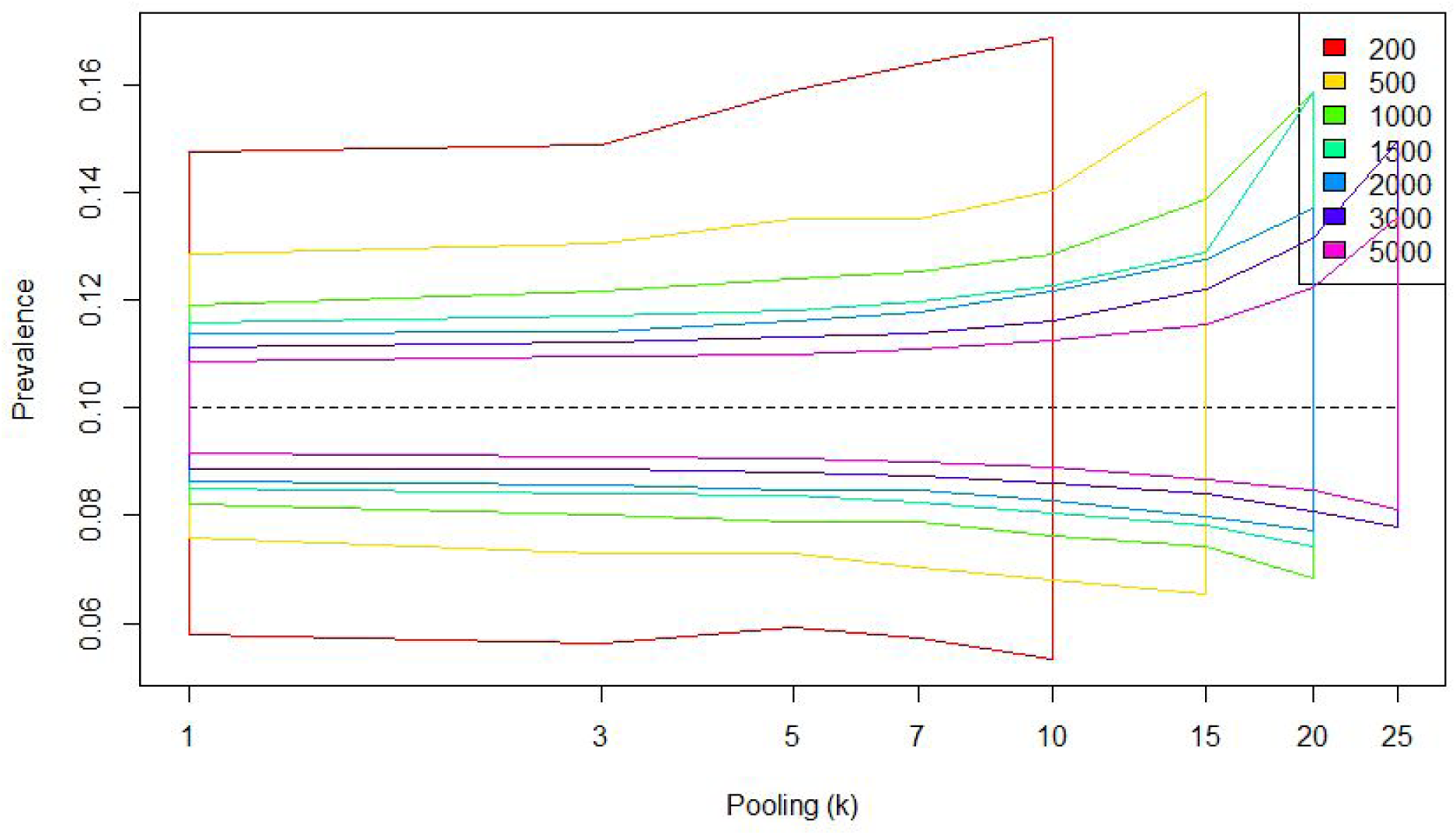
True prevalence = 10%.

### Imperfect specificity

Although there have not been many reports of cross-reactivity between Covid-19 and other viruses in the most commonly used RT-PCR tests, it can be helpful to know how the chance of false positives can impact on the effect of sample pooling. It is also possible that false positives can arise from human errors in the lab. In the following, we repeat the above exercise with specificity (θ) set to 0.99. This creates a seemingly paradoxical situation in which higher levels of sample pooling often leads to more precise prevalence estimates. This is because many, in some cases most, pools are positive without actually containing a single true positive sample, leading to inflated estimates of the prevalence. When the level of pooling goes up, the probability that a positive pool contains at least one true positive sample increases, which increases the total precision.

**Fig. 6.**
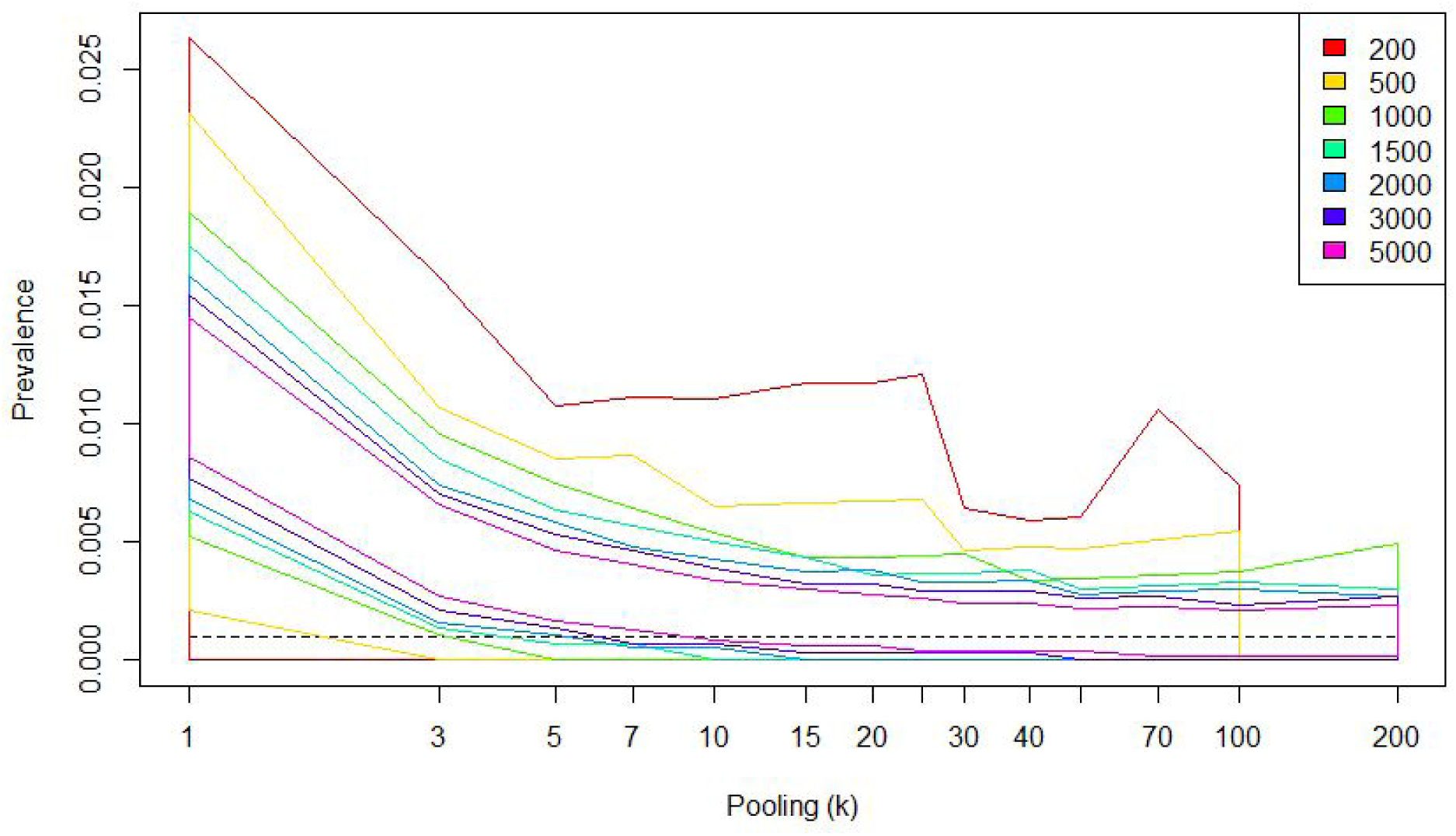
True prevalence = 0.1%, test specificity = 0.99

**Fig. 7.**
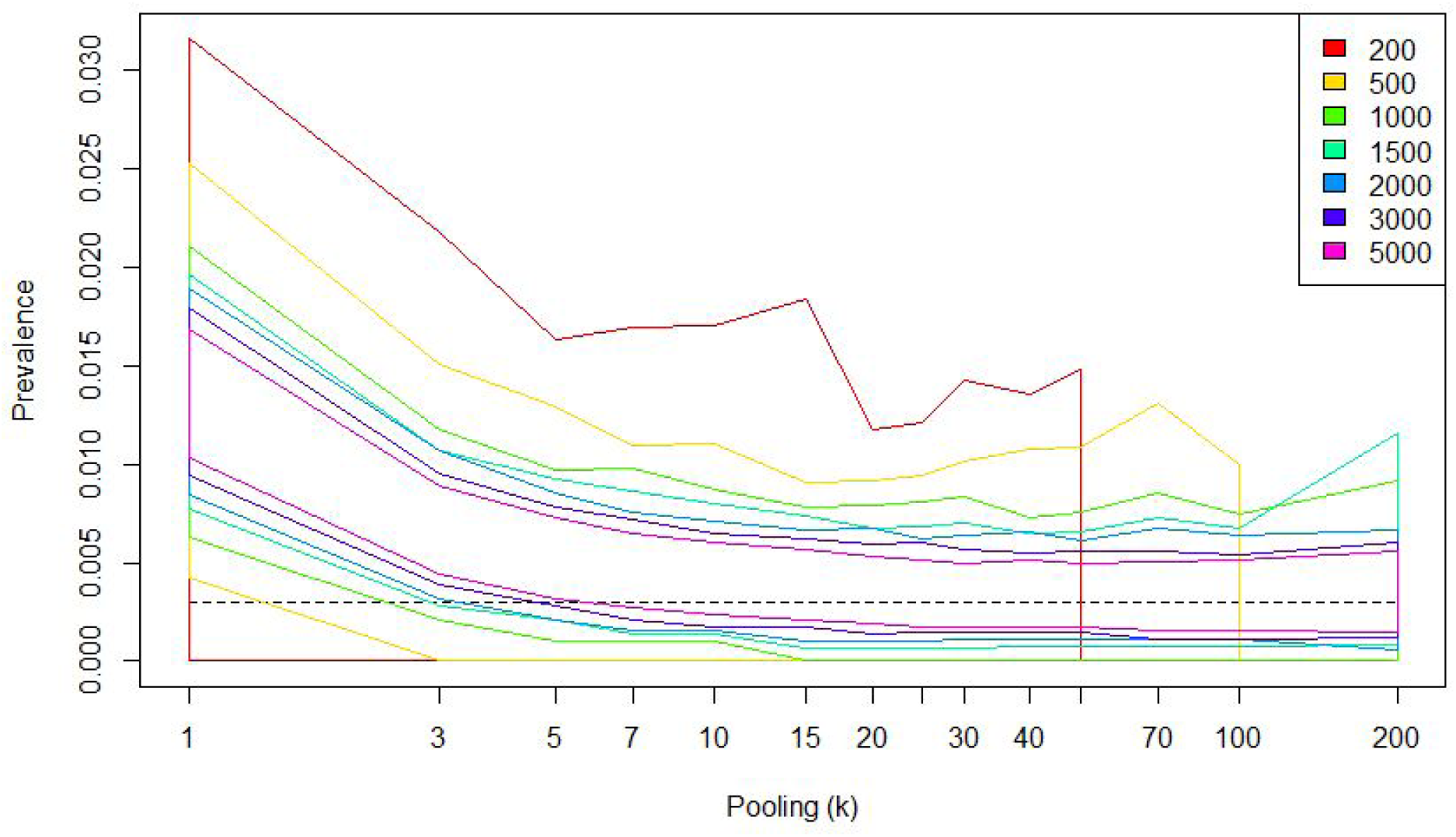
True prevalence = 0.3%, test specificity = 0.99

**Fig. 8.**
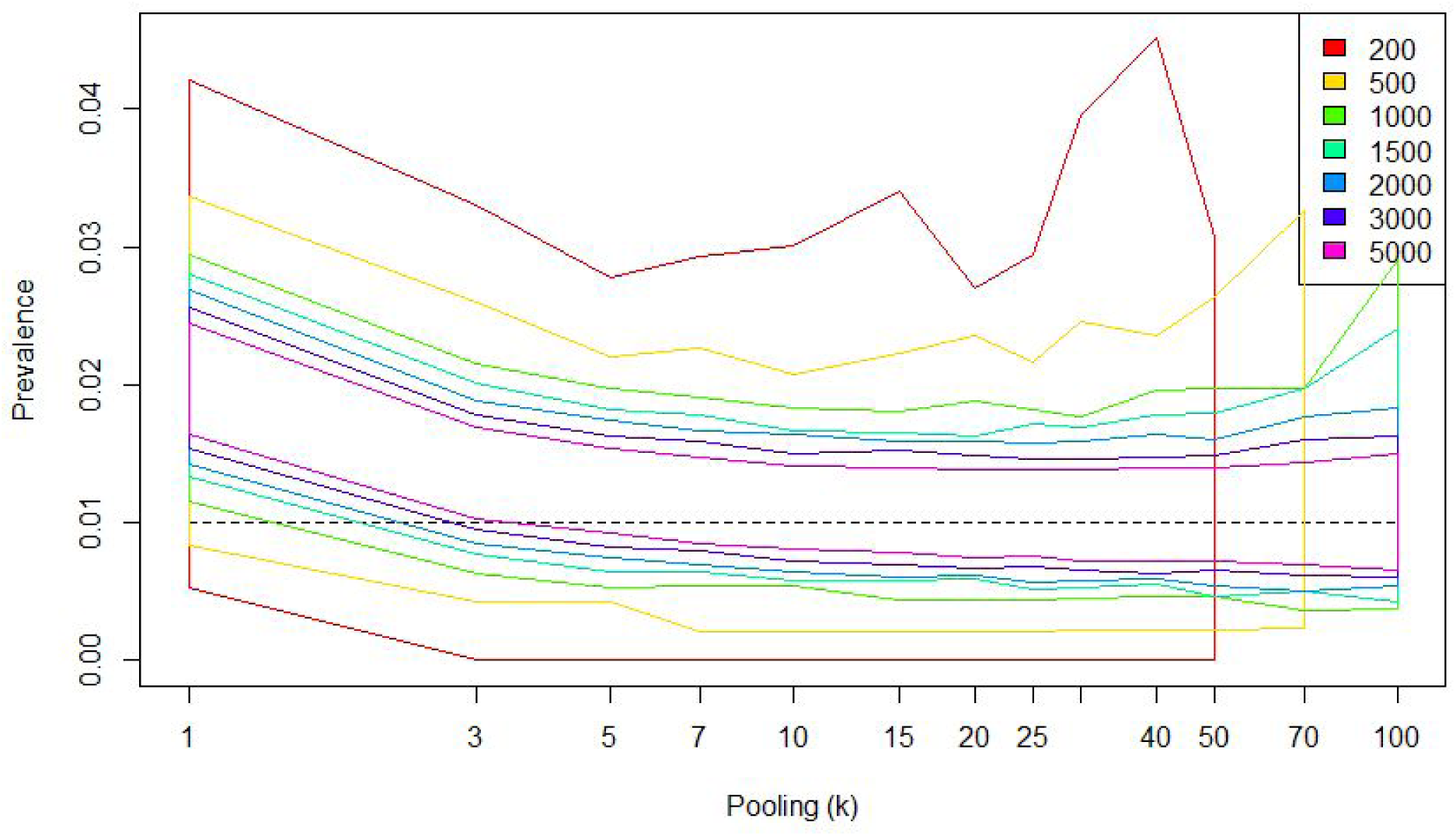
True prevalence = 1%, test specificity = 0.99

**Fig. 9.**
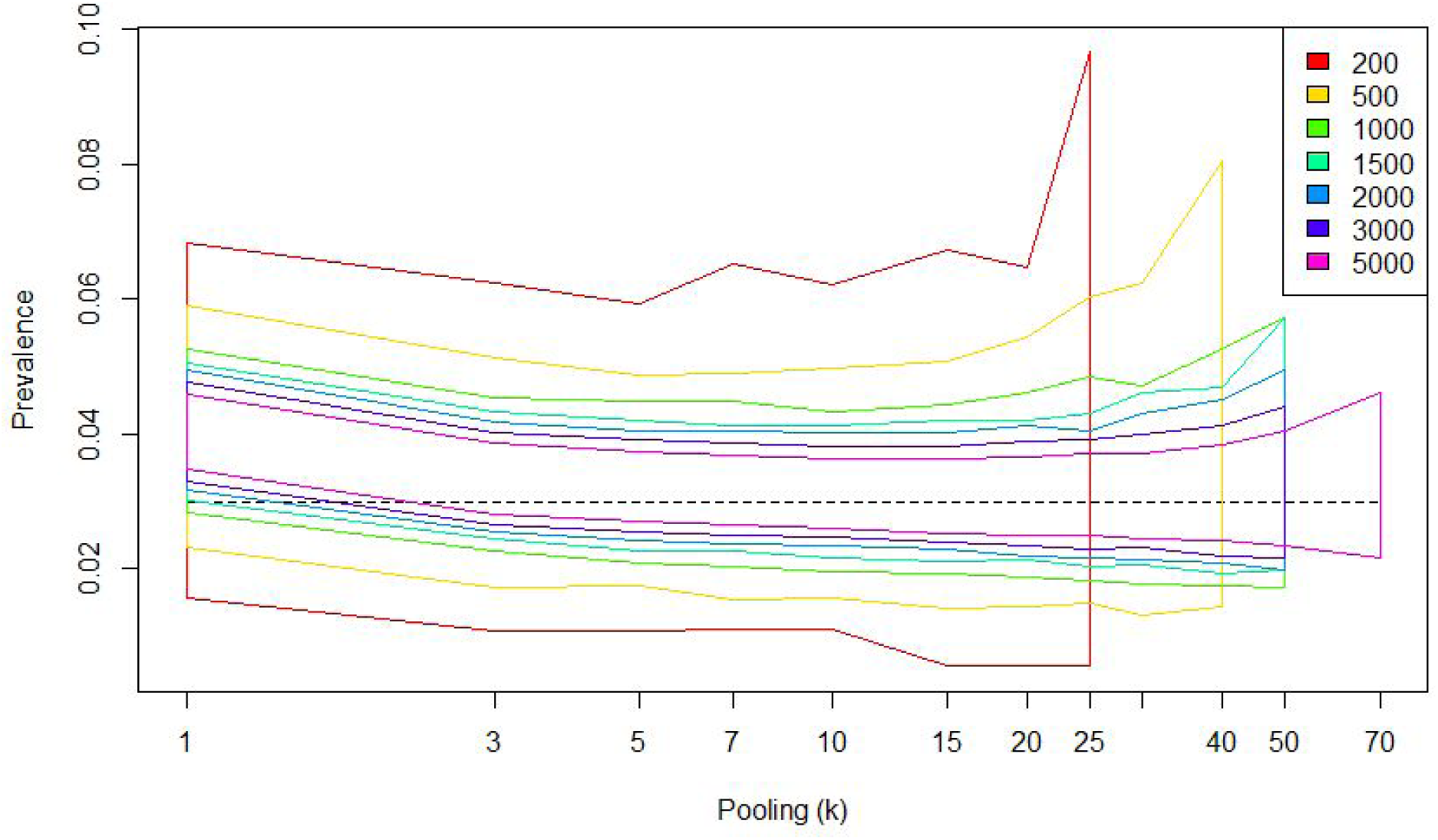
True prevalence = 3%, test specificity = 0.99

**Fig. 10.**
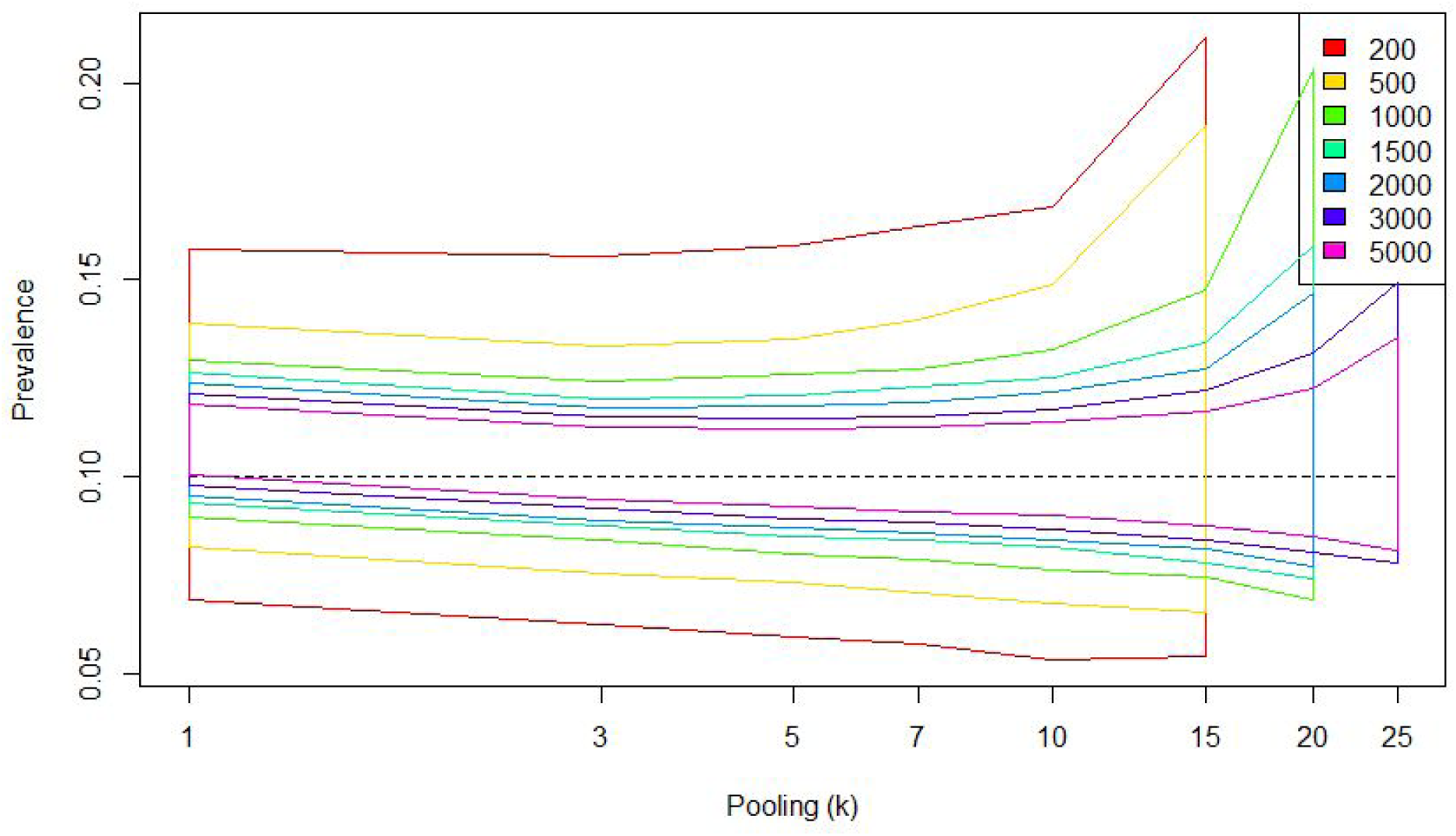
True prevalence = 10%, test specificity = 0.99

### Freedom from disease

Sample pooling can also be of great benefit in order to establish freedom from disease. True freedom in a population is not possible to assert without sampling every last individual. However, we can establish how many samples we need in order to have at least (1 − *α*)% probability of getting a positive sample if the true prevalence was *p*. For example, from figure 6 we can see that if we sample 60 patients and the true prevalence is 0.06, we would be 95% certain that at least one of our samples came out positive. That is, if the true prevalence in the source population was exactly 0.06, we would only expect to get 60 negative samples by chance 5% of the time. A common interpretation of this is that if all pools test negative, we can be 95% certain that the true prevalence in the source population is 0.06 or lower.

Note that when the specificity is 1.0, we will never get a positive from a completely disease-free population no matter how many samples we take. If the test specificity is less than 1.0, the sample size needed to ensure (1 − *α*)% probability of getting at least one positive sample has an upper bound even when the population is free of disease, although this will of course be a false positive.

**Fig. 11.**
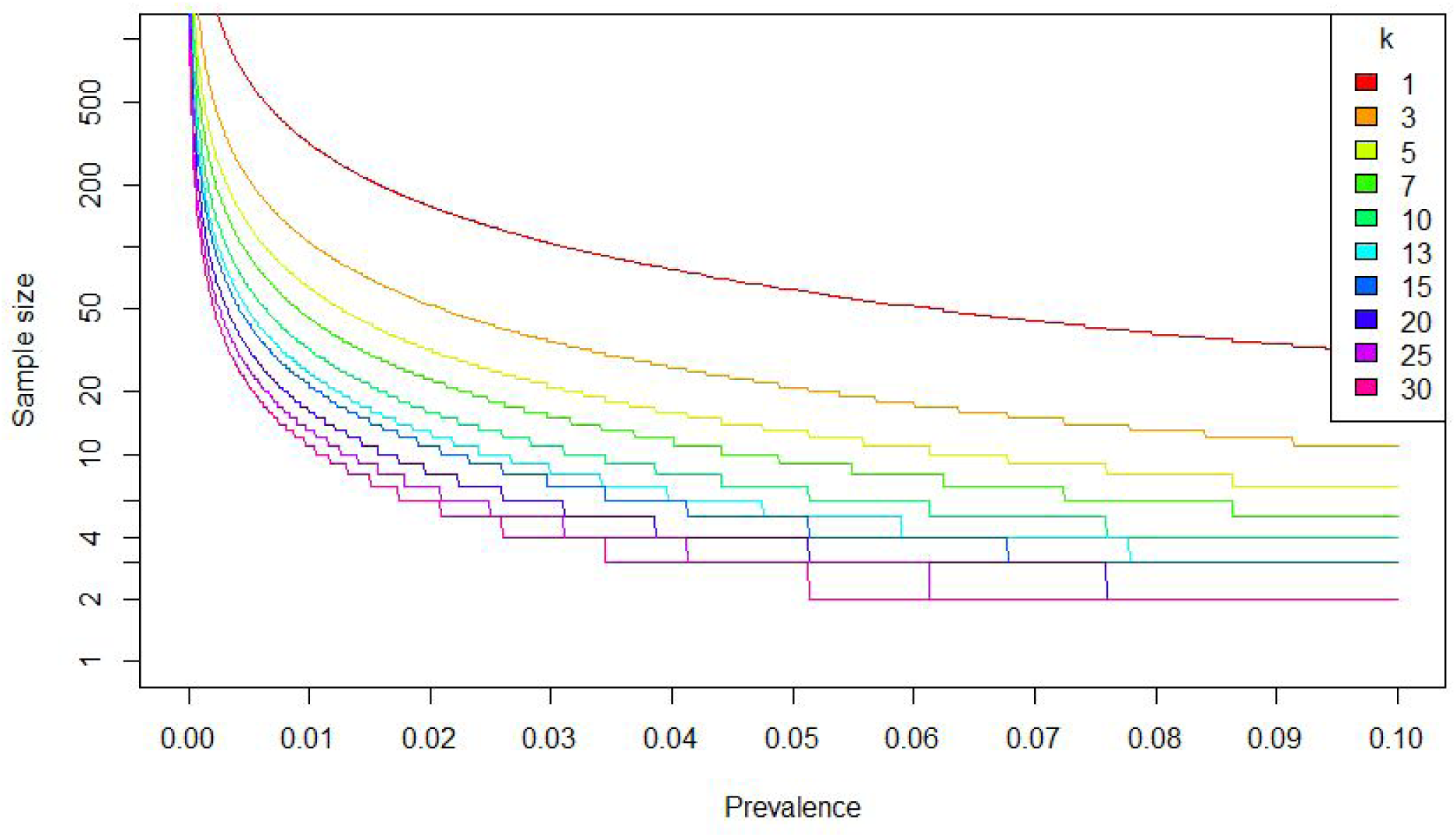
Freedom from disease.

**Fig. 12.**
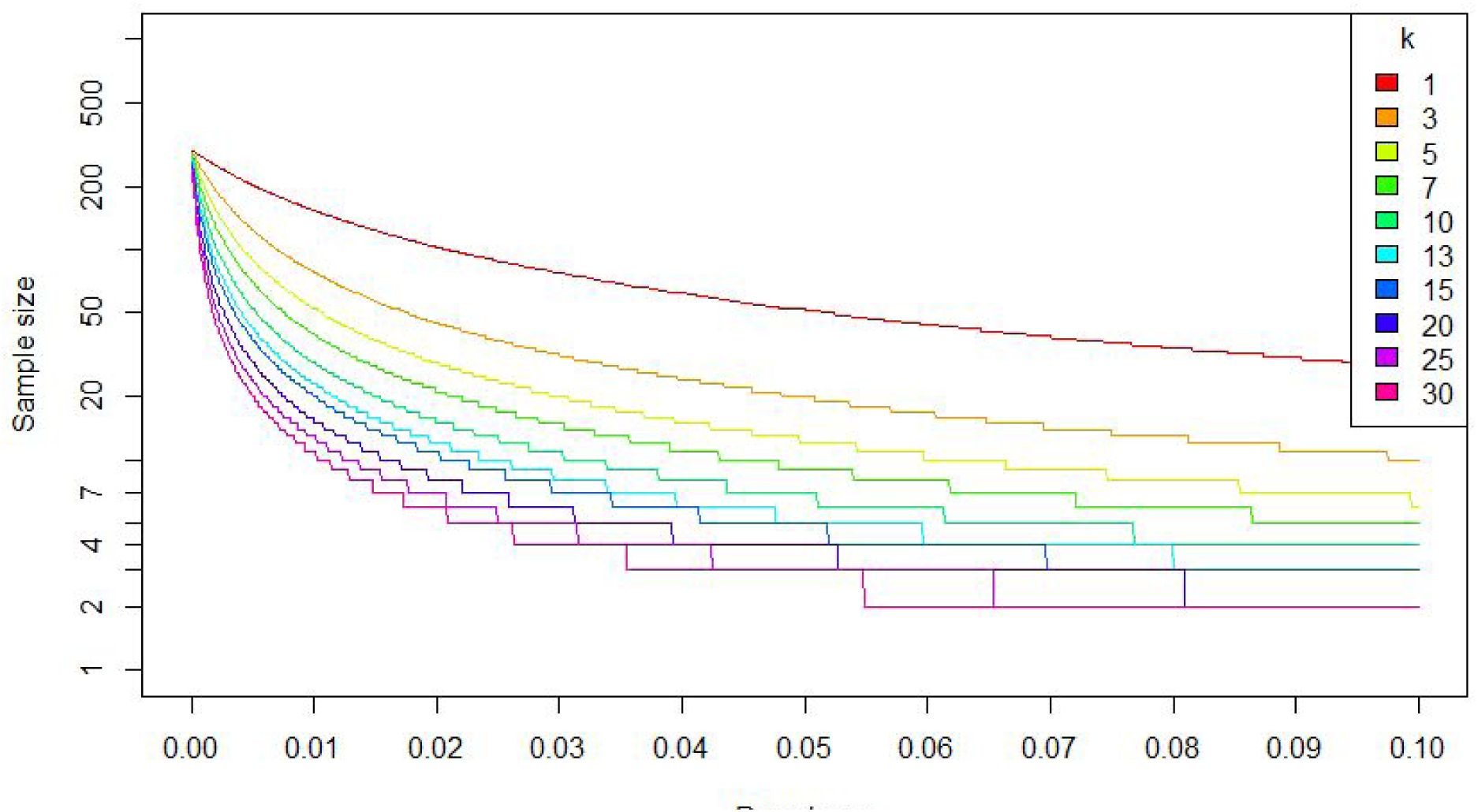
Freedom from disease, specificity 0.99

## SUMMARY

Attempts to estimate the true prevalence of Covid-19 in presumptive low-prevalence or disease-free populations can benefit from sample pooling strategies. Such strategies have the potential to greatly reduce sampling-associated costs with only slight decreases in the precision of prevalence estimates. If the prevalence is low, it is generally appropriate to pool even hundreds of samples, but the total sample count needs to be high in order to get reasonably precise estimates of the true prevalence. On the other hand, if the prevalence is high there is little to be gained by pooling more than 15 samples.

## Data Availability

All data used in this study was generated through simulations. All code is available at https://github.com/admiralenola/pooledsampling-covid-simulation

## AVAILABILITY OF CODE

Code written for this project is available at https://github.com/admiralenola/pooledsampling-covid-simulation. All simulations and plots were created in R version 3.2.3 [8].

